# Dopaminergic stimulants and risk of Parkinson’s disease

**DOI:** 10.1101/2020.05.06.20089748

**Authors:** Michael Wainberg, Dipender Gill, Bowen Su, Mike A. Nalls, Robert R. Graham, Sudeshna Das, Ioanna Tzoulaki, Nasa Sinnott-Armstrong, Manuel A. Rivas

## Abstract

Parkinson’s disease is characterized by dopaminergic neurodegeneration in the substantia nigra. Although dopaminergic drugs are the mainstay of Parkinson’s treatment, their putative disease-modifying properties remain controversial. We explored whether prescription of dopaminergic stimulants for attention-deficit hyperactivity disorder (ADHD) might affect Parkinson’s incidence. We performed Cox survival analyses for outpatient Parkinson’s diagnosis among ADHD-diagnosed seniors in the Optum Clinformatics™ Data Mart de-identified administrative claims database, correcting for diverse demographic and socio-economic status covariates. We compared 5,683 sustained users (≥ 90 days) of dopaminergic stimulants to 252 sustained users of atomoxetine, a noradrenergic first-line ADHD medication. Parkinson’s incidence was reduced among sustained dopaminergic stimulant users compared to atomoxetine users (adjusted hazard ratio [HR] 0.15, 95% confidence interval [CI] 0.04-0.56, p = 0.005). Effect sizes were comparable between derivatives of amphetamine (adjusted HR 0.12, 95% CI 0.03-0.48, p = 0.003) and methylphenidate (adjusted HR 0.27, 95% CI 0.04-1.76, p = 0.2). In sensitivity analyses, similar trends were observed when other psychotropics (SSRIs, gabapentin) were used as comparators instead of atomoxetine, or when the threshold for sustained use was defined as 45, 180 or 360 days instead of 90. Thus, sustained dopaminergic stimulant use was associated with lower Parkinson’s incidence among seniors with ADHD. Our results are consistent with a protective effect of dopaminergic stimulants on the development of Parkinson’s, and support a re-examination of certain dopaminergics, particularly rasagiline and other selective monoamine oxidase B inhibitors, as potential disease-modifying agents.

## Introduction

Parkinson’s disease is one of the most common neurodegenerative diseases, affecting approximately 6 million individuals worldwide, predominantly over age 65^1^. Etiologically, Parkinson’s is characterized by the degeneration of dopaminergic neurons in the substantia nigra and consequent dopamine deficiency in the striatum, leading to motor deficits, mood and sleep disorders, cognitive impairment and hyposmia, among other symptoms^2,3^. Dopaminergic drugs, such as the dopamine precursor L-DOPA; the dopamine agonists pramipexole, ropinirole and rotigotine; and the monoamine oxidase B (MAO-B) inhibitors selegiline and rasagiline effectively treat motor and to a lesser extent non-motor symptoms of Parkinson’s by raising striatal dopamine levels^4^. However, no drug has been approved to delay or reverse the neurodegeneration underlying Parkinson’s disease; such a disease-modifying therapy has been described as “the greatest unmet therapeutic need in Parkinson's disease”^5^.

Though controversial, some evidence suggests that dopaminergic drugs such as rasagiline may influence Parkinson’s-associated neurodegeneration in addition to relieving symptoms. Though the LEAP^6^ and PROUD^7^ delayed-start trials found that early treatment with L-DOPA and pramipexole, respectively, do not significantly alter the rate of progression of Parkinson’s symptoms, the ADAGIO delayed-start trial found that early treatment with rasagiline was associated with significantly reduced rate of progression at 1 mg/day, but not 2 mg/day^8^. The positive result of the ADAGIO trial at 1 mg/day is consistent with experimental evidence that rasagiline^9–16^ and its primary metabolite 1-(R)-aminoindan^15,17^ have neuroprotective properties.

In this study, we aimed to complement these randomized control trials with real-world evidence from an American insurance claims database, Optum Clinformatics™ Data Mart, containing de-identified medical and pharmacy claims for over 57 million unique patients. The uniquely large size of this cohort allowed us to focus on a highly specific patient population, seniors prescribed dopaminergic stimulants (amphetamine and methylphenidate derivatives) for attention-deficit hyperactivity disorder (ADHD). At high doses, amphetamines are neurotoxic via a variety of mechanisms^18^, and mice treated with 10-20 mg/kg of methamphetamine, the equivalent of approximately 50 to 100 times the human therapeutic dose, display ~40-45% dopaminergic neuronal loss in the substantia nigra^19^. Conversely, therapeutic doses of amphetamines have been found to modestly ameliorate Parkinson’s symptoms^20^, and low-dose methamphetamine appears to be neuroprotective in animal models of stroke and traumatic brain injury^21^. Thus, we hypothesized that therapeutic use of dopaminergic stimulants for ADHD might influence Parkinson’s-associated neurodegeneration and therefore the incidence of transition from prodromal to frank disease.

## Methods

### DATA SOURCE

In this cohort study, we used Optum Clinformatics™ Data Mart Database (OptumInsight, Eden Prairie, MN), a de-identified database of medical and pharmacy claims for over 57 million unique patients from a large national U.S. insurance provider.

### STUDY POPULATION

Eligible patients were aged 65 or over at date of cohort entry (see below for definition) and had previously had at least one outpatient encounter with a diagnosis of ADHD (*International Classification of Diseases, 9th Revision* [ICD-9], code 314, or *10th Revision* [ICD-10], code F90). Patients were excluded if they had either no administrative claim entries, outpatient prescription entries, insurance plan entries, or socio-economic status entries in the database. Patients with any entries for dopaminergic stimulants rarely prescribed for ADHD (phentermine, pemoline, and methamphetamine) were excluded, as in a recent insurance claims study of dopaminergic stimulants and psychosis^22^, as were patients with ambiguous ages (different years of birth recorded in different insurance plan entries).

A previous study of dopaminergic stimulants and Parkinson’s in the Utah Population Database found a 50% increased hazard of Parkinson’s among ADHD-diagnosed therapeutic ever-users of stimulants aged 66 and under compared to never-users^23^, but this result could be explained by individuals with missing prescription data being more likely to also lack data on Parkinson’s diagnosis. To avoid this source of confounding in our own study design, we employed a control group consisting of patients taking a comparator drug: atomoxetine, a norepinephrine reuptake inhibitor used, like dopaminergic stimulants, as a first-line ADHD medication.

The cohort entry date was defined as the date on which the patient was prescribed their 90th day of either a dopaminergic stimulant or atomoxetine, not necessarily contiguously. Prescription entries with ≤ 0 days’ supply were discarded. Patients were excluded if they had never been prescribed more than 90 days’ supply of the medication or had ever taken both types of medications. The cohort exit date was either the date the patient developed Parkinson’s or the date of the patient’s last ICD entry in Optum, whichever came first. Patients were excluded if their entry date would fall after their exit date, i.e. if they were prescribed fewer than 90 days’ supply of a dopaminergic stimulant or comparator drug before developing Parkinson’s or attaining their last ICD entry in Optum. Notably, this study design avoids immortal time bias^24^, regardless of the timing of prescriptions prior to the 90-day mark.

The dopaminergic stimulant group consisted of patients prescribed either amphetamines (mixed amphetamine salts, dextroamphetamine and lisdexamfetamine), methylphenidate derivatives (methylphenidate, and dexmethylphenidate), or both types of stimulants. Patients were permitted to switch between stimulants in the same category, and all stimulants in the category were counted towards the 90-day threshold. However, for the amphetamine- and methylphenidate-specific analyses, patients were excluded if they had ever been prescribed both types of stimulants, even outside the entry and exit dates, in order to be conservative.

### STATISTICAL ANALYSIS

For the main analysis, we performed multivariable Cox regression to estimate hazard ratios [HR] and 95% confidence intervals [CI] for Parkinson’s disease onset between the dopaminergic stimulant and comparator groups. Patients were censored if they did not develop Parkinson’s disease before their last ICD entry in Optum. The primary outcome was an ICD code for Parkinson’s disease (ICD-9: 332.0; ICD-10: G20) or a prescription of a medication containing L-DOPA. (Though L-DOPA was historically also prescribed for restless legs syndrome, its use has been largely superseded by gabapentinoids and dopamine agonists for this indication^25^) All analyses were performed using Python 3.6.8 software, using the standard *lifelines* package (version 0.18.4) for Cox regression.

We adjusted for a wide variety of demographic and socio-economic status markers: age at cohort entry (standardized to zero mean and unit variance), as well as the exponential and logarithm of this standardized age to account for potential non-linearity in the relationship between age and Parkinson’s disease even after adjusting for other covariates, sex, race, region of the US, highest degree obtained, home ownership status, household income range, occupation type, number of adults in household, and number of children in household. (For the logarithm of standardized age, standardized age was shifted by the minimum age minus 1 × 10^−10^ so that the argument of the logarithm was positive.) To further mitigate bias, we also included as covariates the fraction of days each patient was prescribed the stimulant or comparator drug during the period between the cohort entry and exit dates (the total days’ supply of all prescriptions during this period, divided by the length of the period), as a proxy for cumulative exposure to the drug over and above the 90-day initial exposure. We also included the total number of prescription entries of any drug during this period, as a proxy for data density or completeness.

To avoid model convergence failure due to collinearity or quasi-complete separation, we discard the rarest value of each categorical covariate when ‘one-hot encoding’ (converting categorical variables to a series of binary numerical variables), and further discard covariates with variance < 1 × 10^−4^ among either the stimulant or comparator groups, or frequency < 10% in the selected cohort.

### DATA AVAILABILITY

Optum Clinformatics™ Data Mart is available by application through the Stanford Center for Population Health Sciences (http://med.stanford.edu/phs.html).

## Results

### STUDY COHORT

The cohort for the primary analysis consisted of 5,683 patients in the dopaminergic stimulants group and 252 patients in the comparator group, with a total of 18,825 person-years of follow-up. Covariate frequencies among the two groups are shown in Table 1.

**Table 1:** Patient covariates and their frequencies. Covariates with frequency <5% in both groups omitted for brevity.

| Variable |  | Dopaminergic<br>stimulants<br>(N = 5,683) | Atomoxetine<br>(N = 252) |
| --- | --- | --- | --- |
| Age (years) |  | 68.60 ± 3.93 | 68.14 ± 3.51 |
| Sex | Female | 58.7% | 59.1% |
|  | Male | 41.2% | 40.5% |
| Race | White | 77.6% | 78.2% |
|  | Unknown | 12.6% | 12.3% |
|  | Black | 5.7% | 6.3% |
| Region | South Atlantic | 28.2% | 29.4% |
|  | East North Central | 15.2% | 16.7% |
|  | Pacific | 11.1% | 11.5% |
|  | Mountain | 12.4% | 7.5% |
|  | West South Central | 8.5% | 10.3% |
|  | West North Central | 7.4% | 8.3% |
|  | Middle Atlantic | 6.5% | 6.7% |
|  | New England | 6.8% | 6.0% |
| Education level | Bachelor's or greater | 22.5% | 27.4% |
|  | Less than bachelor's | 57.5% | 51.2% |
|  | High school diploma | 19.5% | 20.2% |
| Home ownership | Likely homeowner | 71.5% | 69.4% |
|  | Likely non-homeowner | 6.4% | 4.4% |
|  | Unknown | 22.0% | 26.2% |
| Household income<br>range | <\$40K | 19.5% | 20.6% |
| | \$40K-\$49K | 6.4% | 4.4% |
| | \$50K-\$59K | 7.6% | 4.4% |
| | \$60K-\$74K | 10.5% | 9.5% |
| | \$75K-\$99K | 14.4% | 11.1% |
| | \$100K+ | 26.3% | 27.8% |
|  | Unknown | 15.3% | 22.2% |
| Occupation type | Missing/unknown | 75.7% | 73.4% |
|  | White Collar/Health/Civil Service/Military | 9.2% | 13.1% |
|  | Homemaker/Retired | 7.8% | 7.5% |
|  | Manager/Owner/Professional | 5.1% | 4.8% |
| # adults in<br>household | 1 | 88.7% | 76.2% |
|  | 2 | 8.8% | 19.4% |
| # children in<br>household | 0 | 99.7% | 100.0% |

Of the 8,857,739 individuals aged 65 or over with at least one administrative claim entry, outpatient prescription entry, insurance plan entry, and socio-economic status entry in Optum Clinformatics, there were 141,850 (1.6%) cases of Parkinson’s disease according to the definition used for the primary outcome (Methods). There were 40,039 (0.45%) cases of ADHD (the majority of whom did not have a sufficient number of stimulant prescriptions to be included in the dopaminergic stimulant or comparator groups), of which 589 (1.5%) were also cases of Parkinson’s. Thus, ADHD diagnosis was associated with marginally reduced prevalence of Parkinson’s (relative risk 0.92, Fisher p = 0.04) in this dataset.

### PRIMARY ANALYSIS

Of the 5,683 patients in the dopaminergic stimulants group, 14 developed Parkinson’s; of the 252 patients in the atomoxetine group, 3 developed Parkinson’s. Sustained use of dopaminergic stimulants was associated with a lower incidence of Parkinson’s (Table 2), with an adjusted hazard ratio of 0.15 (95% CI, 0.04 to 0.56, p = 0.005). Effect sizes appeared consistent between amphetamines (adjusted hazard ratio 0.12, 95% CI, 0.03 to 0.48, p = 0.003) and methylphenidate derivatives (adjusted hazard ratio 0.27, 95% CI, 0.04 to 1.76, p = 0.2), though with wide confidence intervals on the methylphenidate group.

**Table 2:**
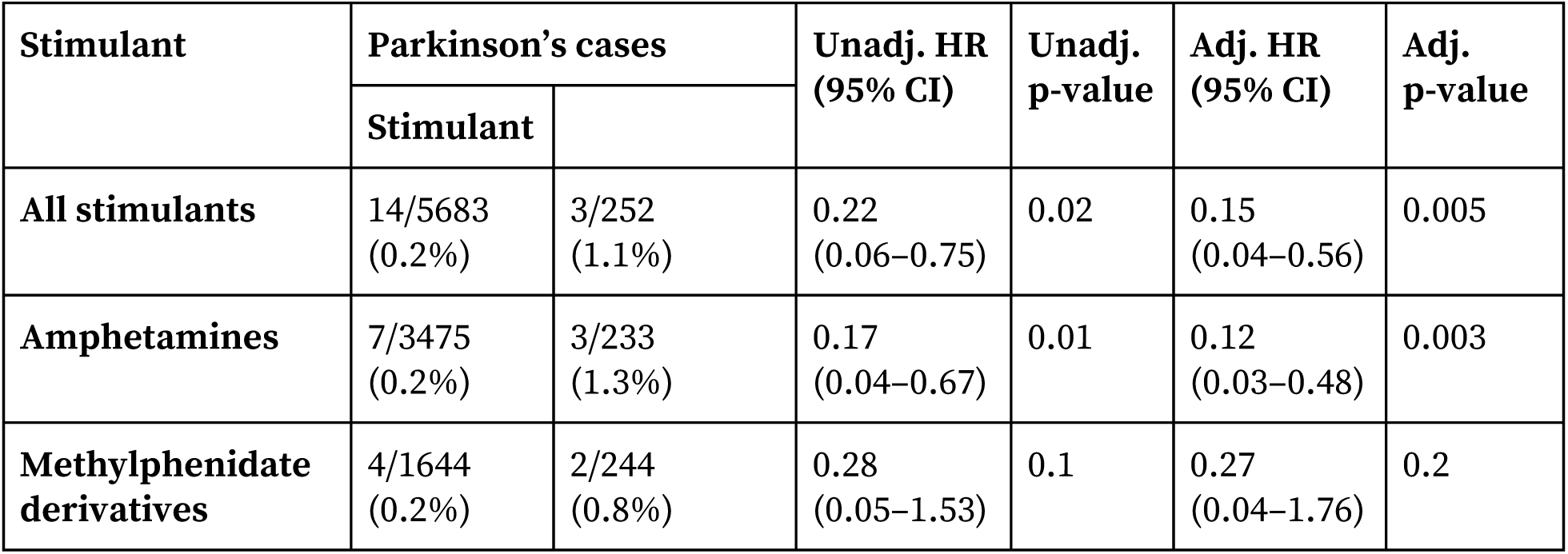
Primary analysis. Unadj. refers to analyses based on raw case incidence; adj. is adjusted for relevant covariates (Methods).

### SENSITIVITY ANALYSES

We performed two types of sensitivity analyses (Table 3). In analyses using either SSRIs (citalopram, escitalopram, fluoxetine, fluvoxamine, paroxetine, and/or sertraline) or gabapentin instead of atomoxetine as the comparator drug (with both the stimulant and comparator groups still restricted to individuals with ADHD), sustained use of dopaminergic stimulants was again associated with a significantly lower incidence of Parkinson’s (SSRIs: adjusted HR 0.36, 95% CI, 0.14 to 0.92, p = 0.03; gabapentin: adjusted HR 0.14, 95% CI, 0.05 to 0.39, p = 0.0001). In analyses using 45, 180 or 360 days as the threshold for sustained use instead of 90, point estimates of the adjusted hazard ratio were similar to the primary analysis (0.10 to 0.34), though sometimes with greater margins of error due to smaller sample sizes.

**Table 3:** Sensitivity analyses. Note that the choice of comparator affects the number of individuals in the stimulant group, due to the exclusion of patients on both the stimulant and the comparator.

| Analysis | Parkinson's cases |  | Unadj. HR<br>(95% CI) | Unadj.<br>p-value | Adj. HR<br>(95% CI) | Adj.<br>p-value |
| --- | --- | --- | --- | --- | --- | --- |
|  | Stimulant | Comparator |  |  |  |  |
| SSRIs as comparator | 8/3295<br>(0.2%) | 14/1845<br>(0.8%) | 0.38<br>(0.16–0.90) | 0.03 | 0.36<br>(0.14–0.92) | 0.03 |
| Gabapentin as comparator | 10/4907<br>(0.2%) | 8/828<br>(1.0%) | 0.19<br>(0.08–0.49) | 0.0005 | 0.14<br>(0.05–0.39) | 0.0001 |
| 45-day threshold | 17/5814<br>(0.3%) | 2/254<br>(0.8%) | 0.38<br>(0.09–1.65) | 0.2 | 0.34<br>(0.08–1.53) | 0.2 |
| 180-day threshold | 12/5165<br>(0.2%) | 4/213<br>(1.9%) | 0.13<br>(0.04–0.40) | 0.0004 | 0.10<br>(0.03–0.32) | 0.0001 |
| 360-day threshold | 11/4058<br>(0.3%) | 2/168<br>(1.2%) | 0.22<br>(0.05–0.98) | 0.05 | 0.25<br>(0.05–1.27) | 0.09 |

### POWER CALCULATION

We performed power calculations according to the formula of Schoenfeld (1983)^26^, using as input the number of individuals in the stimulant and comparator groups, the total number of Parkinson’s cases across both groups, the adjusted hazard ratio, and a significance level of 0.05. Though estimated power was only 47% for the primary analysis due to the small number of seniors prescribed atomoxetine, power was adequate for the sensitivity analyses using larger comparator groups (74% for SSRIs and 90% for gabapentin). The high degree of concordance between the primary and sensitivity analyses suggests the results of the primary analysis are unlikely to be substantially biased due to insufficient power.

## Discussion

Since the dramatic discovery of L-DOPA as a highly effective Parkinson’s disease treatment^27^, it has become well established that dopaminergic drugs are efficacious at controlling the symptoms of the disease. In this study, we add to the body of evidence suggesting that certain dopaminergic drugs may also prevent or delay progression of the neurodegeneration underlying the disease. We find that sustained dopaminergic stimulant use is associated with reduced incidence of Parkinson’s disease among seniors with ADHD.

Our study has several limitations. First, our analysis was restricted to individuals with ADHD. Although this has the benefit of mitigating confounding by indication, our findings in ADHD patients may not fully generalize to individuals without ADHD. Fortunately, a diagnosis of ADHD was on its own associated with only mildly altered Parkinson’s prevalence (relative risk 0.92, Fisher p = 0.04), suggesting that individuals with ADHD are similar to the general population in relation to the development of Parkinson’s disease.

Second, the apparent reduction in Parkinson’s incidence could be driven by dopaminergic stimulants merely masking the symptoms of prodromal disease (and thereby delaying time to diagnosis), without causally affecting neurodegeneration. However, amphetamines were found to only improve Parkinson’s disease symptoms by about 20%^20^, suggesting any masking effect would be unlikely to explain such a large observed reduction in Parkinson’s hazard. Further, dopaminergic stimulants and atomoxetine are rarely prescribed for indications other than ADHD, making it unlikely that reverse causality (patients being prescribed stimulants to treat symptoms of prodromal Parkinson’s disease) could explain the observed results.

Third, the limited number of years of available data (only since 2003) and the inherent incompleteness of insurance claims mean that some individuals could have been diagnosed with ADHD and started on stimulants long before their cohort entry date. Indeed, though ADHD is increasingly being diagnosed in adults and adult-onset ADHD has even been postulated to be a distinct disorder from child-onset ADHD^28^, some individuals in our cohort may well have been taking stimulants for ADHD since childhood. While his “tip of the iceberg effect”, in which the small number of stimulant prescriptions visible in Optum are a marker for a potentially much larger number of unobserved prescriptions, helps to justify the large effect sizes we observe, it could also introduce subtle biases that are difficult to fully account for (though not immortal time bias - see Methods). For instance, some individuals in the atomoxetine group could have originally been treated with dopaminergic stimulants prior to atomoxetine’s FDA approval in 2002, though if anything this would tend to make dopaminergic stimulants look less beneficial, rather than more beneficial.

Fourth, though our study supports dopaminergics being disease-modifying in prodromal PD, this does not necessarily imply being disease-modifying in established PD. This distinction is supported by the relative lack of overlap in genetic risk factors between Parkinson’s risk and Parkinson’s progression found by genome-wide association studies^29^.

Fifth, the low rates of recorded ADHD diagnosis and stimulant use among seniors, combined with the relative rarity of Parkinson’s disease, limit the power of our analysis; and, as with all observational data analysis, our observed association may not be causal. For instance, though observational studies linked high levels of urate (an antioxidant) to both reduced risk of Parkinson’s and slower disease progression, the Study of Urate Elevation in Parkinson's Disease (SURE-PD3) trial did not show disease-modifying effects from raising urate levels with inosine supplementation. We emphasize the need for replication in other cohorts (though very few, if any, others exist of a suitable size), and above all randomized control trial evidence, to validate the conclusions presented here.

Although the ADAGIO trial has sometimes been perceived as blanket evidence against the neuroprotective efficacy of rasagiline in Parkinson’s, with one commentary^30^ stating categorically that “the earliest suggestions of any [disease-modifying] effect for […] rasagiline in TEMPO [a non-delayed-start trial] were negated by the rigorous subsequent delayed-start [trial] ADAGIO”, this glosses over the substantial complexity and ambiguity of the trial’s results. Perhaps the most parsimonious interpretation of the ADAGIO trial is that the delayed-start arms did suffer from greater neurodegeneration, but that the larger 2 mg/day dose was sufficient to mask the additional symptoms resulting from this neurodegeneration, while the smaller 1 mg/day dose was not. Crucial support for this interpretation is provided by the observation in the ADAGIO paper that “for rasagiline at a dose of 2 mg, a post hoc subgroup analysis showed that for subjects in the highest quartile of UPDRS [Unified Parkinson's Disease Rating Scale] scores at baseline, early-start rasagiline provided a significant benefit over delayed-start rasagiline with respect to the change in the UPDRS score between baseline and 72 weeks (−3.63 UPDRS points), and all primary end points were met despite the relatively small sample”. The notion that dopaminergic drugs as a class are either all disease-modifying or all non-disease-modifying may likewise be overly simplistic: it is entirely possible that rasagiline is disease-modifying as a result of its demonstrated neuroprotective effects^9–16^, while L-DOPA is not as a result of neurotoxic effects^31^ negating neuroprotective effects. Our results suggest that the ADAGIO trial’s finding of delayed disease progression with 1 mg/day rasagiline should not be so easily dismissed, and support further randomized clinical trials of rasagiline (or alternatively of even more selective MAO-B inhibitors such as sembragiline^32^) to resolve the ambiguity and either replicate or definitively refute this finding. Future trial designs could account for any potential masking effect by including a washout phase at the end of the trial in addition to a delayed-start phase at the beginning of the trial.

In conclusion, sustained dopaminergic stimulant use was associated with reduced incidence of Parkinson’s disease among seniors with ADHD. Given the urgent unmet need for disease-modifying therapies in Parkinson’s, our results support further investigation of the use of selective monoamine oxidase B inhibitors, and possibly other dopaminergics, as potential disease-modifying agents in Parkinson’s.

## Acknowledgments

The authors would like to thank Rocky Aikens, Deborah Blacker, Mark Albers and John Hsu for helpful discussions. This work was supported by a Stanford Bio-X Bowes Fellowship (to M.W.), Stanford Graduate Fellowship (to N.S.-A.), and National Defense Science & Engineering Graduate Fellowship (to N.S.-A.). M.A.N.’s participation in this work was supported in part by a consulting contract between Data Tecnica International and the National Institute on Aging.

Ethics approval and data access for this project were obtained through the Stanford Center for Population Health Sciences. The PHS Data Core is supported by a National Institutes of Health National Center for Advancing Translational Science Clinical and Translational Science Award (UL1 TR001085) and internal Stanford funding. M.A.R. is supported by Stanford University, a National Institute of Health center for Multi- and Trans-ethnic Mapping of Mendelian and Complex Diseases grant (5U01 HG009080), and the National Human Genome Research Institute of the National Institutes of Health under Award Number R01HG010140. The content is solely the authors’ responsibility and does not necessarily represent the official views of the NIH.

## Author contributions

M.W., N.S.-A. and M.A.R. conceived of the study. M.A.R. and N.S.-A. supervised the study. M.W., D.G and B.S. performed analyses. M.A.N., R.R.G, S.D. and I.T. provided key conceptual support. M.W. wrote the manuscript with assistance from all authors.

